# Omicron neutralizing antibody response following booster vaccination compared with breakthrough infection

**DOI:** 10.1101/2022.04.11.22273694

**Authors:** Marcel E. Curlin, Timothy A. Bates, Gaelen Guzman, Devin Schoen, Savannah K. McBride, Samuel D. Carpenter, Fikadu G. Tafesse

## Abstract

The rapid spread of the vaccine-resistant Omicron variant of SARS-CoV-2 presents a renewed threat to both unvaccinated and fully vaccinated individuals, and accelerated booster vaccination campaigns are underway to mitigate the ongoing wave of Omicron cases. The degree of immunity provided by standard vaccine regimens, boosted regimens, and immune responses elicited by the combination of vaccination and natural infection remain incompletely understood. The relative magnitude, quality and durability of serological responses, and the likelihood of neutralizing protection against future SARS-CoV-2 variants following these modes of exposure are unknown but are critical to the future trajectory of the COVID-19 pandemic. In this study of 99 vaccinated adults, we find that compared with responses after two doses of an mRNA regimen, the immune responses three months after a third vaccine dose and one month after breakthrough infection due to prior variants show dramatic increases in magnitude, potency, and breadth, including increased antibody dependent cellular phagocytosis and robust neutralization of the recently circulating Omicron variant. These results suggest that as the number of Omicron cases rise and as global vaccination and booster campaigns continue, an increasing proportion of the world’s population will acquire potent immune responses that may be protective against future SARS-CoV-2 variants.

## INTRODUCTION

Since 2020, the global COVID-19 pandemic has been punctuated by episodic waves of increased incidence associated with the emergence of new SARS-CoV-2 variants with progressively greater transmissibility and resistance to immune responses elicited by currently approved vaccines. The most epidemiologically important variants have been classified as variants of concern (VOC) by the World Health Organization, and include Alpha, Beta, Gamma, Delta, and Omicron. Presently, the Omicron variant (including sub-linages BA.1 and BA.2) is the globally dominant circulating strain, notable for its high transmissibility and resistance to neutralization by vaccine-induced antibodies (1,2). The spike protein of the Omicron variant contains 39 amino acid changes (30 substitutions, 6 insertions, and 3 deletions), nearly half of which fall within the receptor binding domain (RBD) that is responsible for binding to the human receptor angiotensin converting enzyme 2 (ACE2) (3). All known neutralizing antibodies bind to the spike protein, with the vast majority targeting the RBD (4–6). Mutations within this region have caused a dramatic loss of neutralizing ability among several of the approved monoclonal antibody therapeutics, and has resulted in restriction of their use in cases of infection with Omicron (7).

It is known that the additional antigenic exposure from boosters and breakthrough infections bolster serological immunity, and third-dose vaccine booster campaigns are underway worldwide to mitigate the ongoing wave of omicron cases (8,9). Vaccine breakthrough infections can directly train the immune system against variant spike proteins, but come with medical risks including prolonged illness (long COVID) and death (10,11). Conversely, booster vaccination is generally safe, and has been shown to effectively increase the neutralizing response against Omicron (12–14). The durability of responses due to boosting and breakthrough infection are unknown, but antibody levels have been shown to decrease over time following primary vaccination, suggesting that waning of the augmented immunity following additional exposure is likely (15,16). It is also unknown whether recovery from breakthrough infection or booster vaccination provide greater protection from reinfection with Omicron and future variants, which will likely affect the future trajectory of the pandemic. To address these knowledge gaps, we examined serological immune responses and antibody-dependent cell-mediated phagocytosis in individuals who had received either two doses of a standard vaccine regimen, a standard regimen followed by a booster, or breakthrough infection following vaccination.

## METHODS

### Cohort selection and serum collection

Two- and three-dose group participants were selected from a larger cohort of vaccinated health care workers at Oregon Health & Science University recruited at the time of their first vaccine dose. Participants were asked to return after either their second or third vaccine dose to provide whole blood samples. Breakthrough group participants were recruited and enrolled at Oregon Health & Science University from among fully vaccinated health care workers receiving positive results during PCR-based diagnostic testing for SARS-CoV-2 infection, at which time participants provided information on symptoms of illness by direct interview. Whole blood (4-6 mL) was collected with a BD Vacutainer® Plus Plastic Serum Tube and centrifuged for 10 minutes at 1000xg, then stored at −20°C. Two- and three-dose group participants confirmed no history of COVID-19 by direct interview and validated by nonreactivity in a SARS-CoV-2 N protein ELISA. All vaccines used in this study were BNT162b2 (Pfizer) and only individuals with no reported immunocompromising conditions were included.

### Enzyme-linked immunosorbent assays (ELISA)

ELISAs were performed as previously described.^1^ In 96-well plates (Corning Incorporated, EIA/RIA High binding, Ref #359096). Plates were coated with SARS-CoV-2 RBD (produced in Expi293F cells and purified using Ni-NTA chromatography), N (SARS-CoV-2 Nucleocapsid-His, insect cell-expressed, SinoBio Cat: 40588-V08B, Item #NR-53797, lot #MF14DE1611) at 100 µL/well at 1 µg/mL in PBS and incubated overnight at 4°C with rocking. Plates were washed three times with .05% Tween-20 in PBS (wash buffer) and blocked with 150 uL/well with 5% nonfat dry milk powder and .05% Tween 20 in PBS (blocking buffer) at room temperature (RT) for 1 hour with rocking. Breakthrough and control sera were aliquoted and frozen in dilution plates then resuspended in blocking buffer; sera were diluted and added to ELISA plates 100 µL/well (6 × 4-fold dilutions from 1:50 to 1:51,200, except for IgM (6 × 4-fold dilutions from 1:25 to 25,600). Sera was incubated in coated plates for 1 hour at RT, then washed three times with wash buffer. Plates were incubated with anti-human IgA-HRP at 1:3,000 (BioLegend, Ref #411002), Mouse anti-human IgG-HRP Clone G18-145 at 1:3,000 (BD Biosciences, Ref #555788), or Goat anti-human IgM-HRP at 1:3,000 (Bethyl Laboratories, Ref #A80-100P) at RT for 1 hour with rocking, then washed three times with wash buffer prior to developing with o-phenylenediamine dihydrochloride (OPD, Thermo Scientific #34005) according to manufacturer instructions. The reaction was stopped after 25 minutes using an equivalent volume of 1 M HCl; optical density was measured at 492 nm using a CLARIOstar plate reader.

### Antibody dependent cellular phagocytosis (ADCP)

ADCP was assessed as described previously.^2^ Biotinylated SARS-CoV-2 RBD protein was incubated with neutravidin beads (Invitrogen, F8775) for 2 hours at room temperature then washed with PBS with 1% BSA (dilution buffer) two times. 10 µL of 1:100 diluted RBD beads were incubated with an equal volume of diluted serum for 2 hours at 37°C. Bead-serum mixtures were then incubated with 20,000 THP-1 cells in a final volume of 100 uL overnight in a tissue culture incubator. 100 µL of PBS with 4% formaldehyde was then used to fix each well for 30 minutes prior to flow cytometry. Triplicate, samples were flowed on a CytoFLEX flow cytometer. 2500 events were recorded per replicate. Phagocytosis scores were calculated as the product of percent bead-positive cells and mean fluorescence intensity of bead-positive cells, then divided by 10^6^.

### SARS-CoV-2 growth and titration

SARS-CoV-2 isolates USA-WA1/2020 [lineage A] (NR-52281), hCoV-19/USA/PHC658/2021 [lineage B.1.617.2 – Delta] (NR-55611), and hCoV-19/USA/MD-HP20874/2021 [lineage B.1.1.529 – Omicron] (NR-56461) were obtained from BEI Resources. Viral stocks were propagated as previously described.^3^ Sub-confluent Vero E6 cells (CRL-1586) grown in Dulbecco’s Modified Eagle Medium (DMEM), 10% fetal bovine serum (FBS), 1% nonessential amino acids, 1% penicillin-streptomycin (complete media) were infected at an MOI of 0.05 in a minimal volume (0.01 mL/cm^2^) of Opti-MEM + 2% FBS (dilution media) for 1 hour at TCC then 0.1 mL/cm^2^ additional complete media was added and incubated until at least 20% cytopathic effect (CPE) was observed, typically 72-96 hours. Culture supernatant was centrifuged for 10 min at 1000xg and frozen at −80°C. Titration was performed by focus forming assay on sub-confluent Vero E6 cells. 10-fold dilutions were prepared in dilution media and incubated for 1 hour, then covered with Opti-MEM, 2% FBS, 1% methylcellulose (overlay media) and incubated for 24 hours (48 hours for Omicron). Plates were then fixed in 4% formaldehyde in phosphate buffered saline (PBS) for 1 hour then removed from BSL3 following institutional guidelines. Cells were permeabilized in 0.1% bovine serum albumin (BSA), 0.1% saponin in PBS (perm buffer) for 30 minutes, then with polyclonal anti-SARS-CoV-2 alpaca serum (Capralogics Inc.) (1:5000 in perm buffer, or 1:2000 for Omicron) overnight at 4°C. Plates were washed three times with 0.01% Tween 20 in PBS (wash buffer), then incubated for 2 hours at RT with 1:20,000 anti-alpaca-HRP (Novus #NB7242), or 1:5000 for Omicron. Plates were washed three times with wash buffer, then incubated with TrueBlue (Sera Care #5510-0030) for 30 minutes or until sufficiently developed for imaging. Foci images were captures with a CTL Immunospot Analyzer and counted with Viridot (1.0) in R (3.6.3).^4^ Viral stock titers in focus forming units (FFU) were calculated based on the dilution factor and volume used for infection.

### Focus reduction neutralization test (FRNT)

FRNT assays were carried out as previously described.^3^ We prepared 5×4.7-fold (1:10-1:4879) serial dilutions in duplicate for each serum sample. An equal volume of viral stock was added to each well (final dilutions of sera, 1:20 – 1:9760) such that approximately 50 FFU were added to each well. Virus-serum mixtures were incubated for 1 h before being used to infect sub-confluent Vero E6 cells in 96-well plates for 1 hour, then covering with 150 µL/well overlay media. Each 5-point serum dilution series was accompanied by a virus only control well. Fixation, development, and counting of FRNT plates was carried out as described in SARS-CoV-2 growth and titration. Percent neutralization values were calculated for each well as focus count divided by the average of virus-only wells from the same plate.

### Statistical analysis

FRNT_50_ and EC_50_ values were calculated by fitting to a dose-response curve as previously described.^3^ Final FRNT_50_ values below the limit of detection (1:20) were set to 1:19. Final EC_50_ values below the limit of detection of 1:25 for N, full-length Spike, Spike RBD, IgG, IgA were set to 1:24 and 1:12.5 for IgM was set to 1:12. Aggregated EC_50_ and FRNT_50_ values were analyzed in Graphpad Prism (9.3.1). Significance was determined using Kruskal-Wallis tests with Dunn’s multiple comparison correction, P-values were two-tailed. Correlations were calculated with log-transformed EC_50_ and/or FRNT_50_ values with the Spearman method, with corresponding two-tailed P values. Best fit lines were calculated via simple linear regression.

### Study approval

This study was conducted with approval of the Oregon Health and Sciences University Institutional review board (IRB# 00022511). All participants were enrolled following written informed consent.

## RESULTS

### Cohort

A total of 97 individuals were studied (Table 1). Participants from the two-dose group provided serum samples a median of 24 days after the second dose. The three-dose group received a third vaccine dose a median of 253 days after the second, and then provided serum samples a median of 86 days after the third dose. Both two- and three-dose groups reported no history of SARS-CoV-2 infection and displayed a lack of Nucleocapsid antibodies (Fig. 1). Breakthrough group participants received positive PCR-based COVID-19 test results a median of 159.5 days after their final vaccine dose and provided serum samples a median of 29 days after the date of PCR testing (Table 1 and Figure 2A).

**Table 1:** Cohort demographics and clinical data

| Characteristic | Two-dose | Three-dose | Breakthrough |
| --- | --- | --- | --- |
| **Cohort characteristics** |  |  |  |
| N | 42 | 27 | 30 |
| Female - N (%) | 35 (83.3) | 19 (70.4) | 23 (76.7) |
| Male - N (%) | 7 (16.7) | 8 (29.6) | 7 (23.3) |
| Age (yr) | 40 [23-74] | 47 [26-74] | 38 [24-63] |
| **Time intervals (days)** |  |  |  |
| Last vaccine to blood draw | 24 [17.25-35.75] | 86 [79.5-93.5] | N/A |
| PCR positivity to blood draw | N/A | N/A | 29 [23-47] |
| 2nd vaccine to positive PCR | N/A | N/A | 159.5 [81.25-202.25] |
| Time between 1st and 2nd vaccines | 21 [21-22] | 21 [21-22] | 21 [21-23] |
| Time between 2nd and 3rd vaccines | N/A | 253 [249-263.5] | N/A |
| **Vaccine type** |  |  |  |
| BNT162b2 (Pfizer) | 42 (100) | 27 (100) | 28 (93.3) |
| mRNA-1237 (Moderna) | 0 (0) | 0 (0) | 1 (3.3) |
| Ad26.COV2.S (Janssen) | 0 (0) | 0 (0) | 1 (3.3) |
| **Breakthrough infection strain** |  |  |  |
| Alpha (B.1.1.7) | N/A | N/A | 4 (16.7) |
| Beta (B.1.351) | N/A | N/A | 1 (3.3) |
| Gamma (P.1) | N/A | N/A | 3 (10) |
| Delta (B.1.617.2) | N/A | N/A | 10 (33.3) |
| Unknown | N/A | N/A | 11 (36.7) |

**Figure 1:**
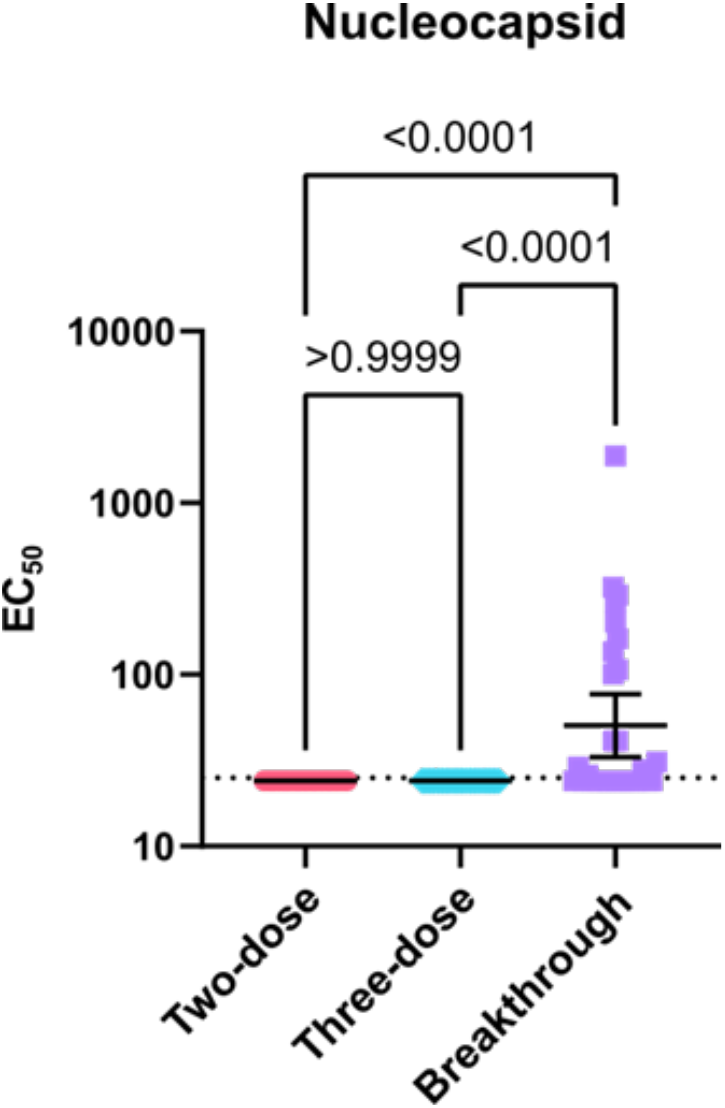
Nucleocapsid ELISA. Serum dilutions with half-maximal binding (EC_50_) of IgG/A/M antibodies to SARS-CoV-2 Nucleocapsid protein. Error bars indicate the geometric mean and 95% confidence intervals. P-values show the results of a two-tailed Kruskal-Wallis test with Dunn’s multiple comparison correction.

**Figure 2:**
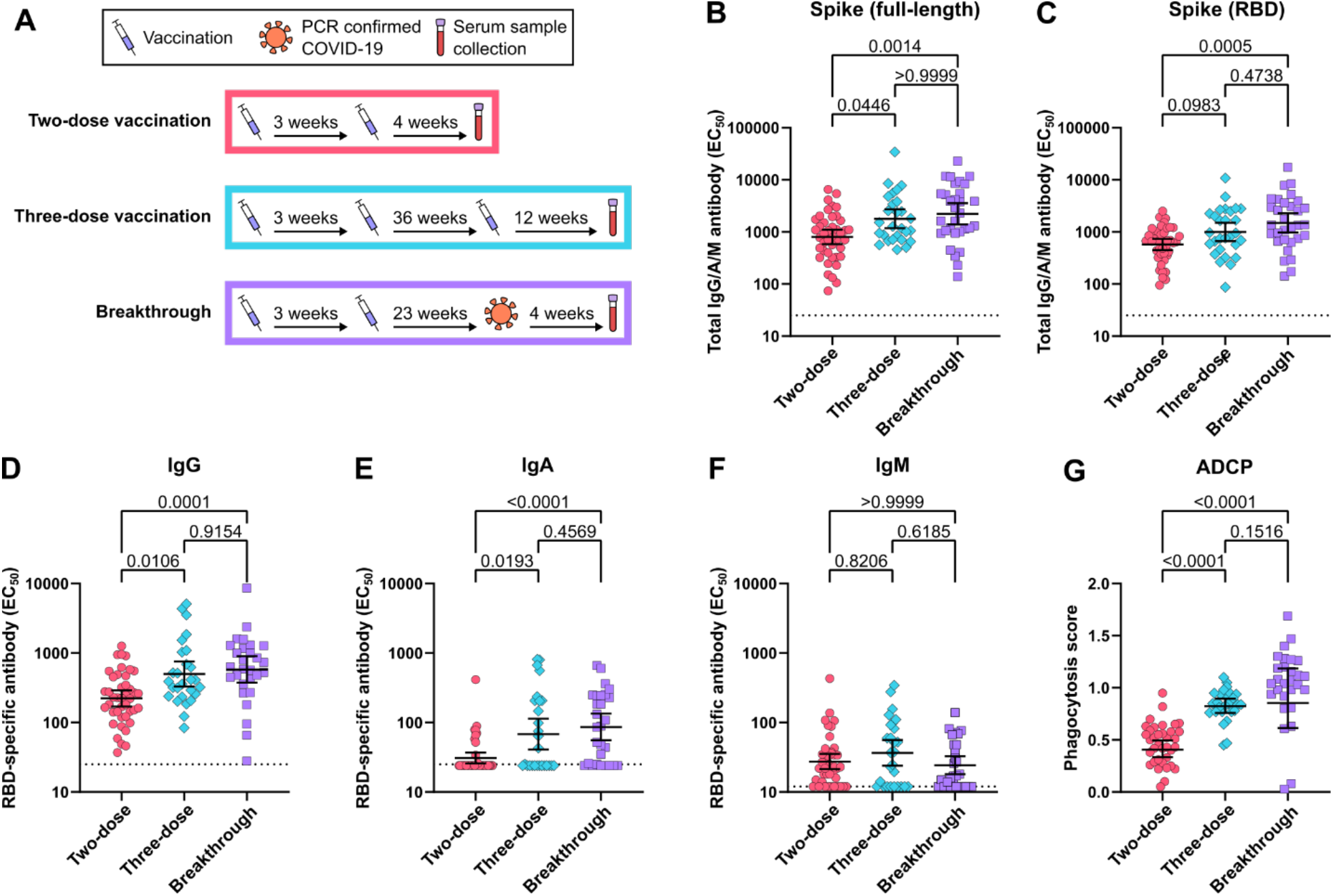
Antibody response to two-dose vaccination, three-dose vaccination and breakthrough infection. (A) Schematic describing median cohort vaccine dose, PCR-confirmed natural infection, and sample collection timing. (B) Serum dilutions with half-maximal binding (EC_50_) of IgG/A/M antibodies to full-length SARS-CoV-2 spike protein. (C) Serum IgG/A/M antibody EC_50_ to spike receptor binding domain (RBD). Serum (D) IgG-specific, (E) IgA-specific, (F) IgM-specific antibody EC50’s to RBD. (G) Antibody dependent cellular phagocytosis scores indicate the increase in uptake of RBD-coated beads caused by sera. Error bars in B-G indicate the geometric mean and 95% confidence intervals. P-values in B-G show the results of two-tailed Kruskal-Wallis tests with Dunn’s multiple comparison correction.

### Approach

In each sample, we analyzed the spike receptor-binding domain-specific IgG, IgA, and IgM antibody levels by enzyme-linked immunosorbent assay (ELISA). We also measured each serum’s ability to neutralize authentic wild-type SARS-CoV-2 (WA1, Wuhan strain) and clinical isolates of the Delta (B.1.617.2) and Omicron (B.1.1.529) variants with focus reduction neutralization tests (FRNT). Finally, we examined the ability of serum in each group to trigger antibody-depended cell-mediated phagocytosis (ADCP) of spike protein-coated beads.

### Binding antibody responses

Compared to two-dose vaccination, the geometric mean of serum dilutions with half-maximal binding in ELISA (EC_50_) to full-length SARS-CoV-2 spike protein was 223% higher in the three-dose group and 278% higher in the breakthrough group; the three-dose and breakthrough groups were not significantly different from each other (Fig. 2B). Spike RBD specific antibodies did not significantly increase in the three-dose group but did in the breakthrough group, which was 259% higher than in the two-dose group (Fig. 2C). Spike-specific IgG and IgA levels showed similar increases relative to the two-dose group, with 224% and 261% higher IgG levels and 221% and 279% higher IgA in the three-dose and breakthrough groups, respectively (Fig. 2D and 2E). IgM levels were not significantly different between any of the groups (Fig. 2F).

### Antibody-dependent cell-mediated phagocytosis

Similarly to neutralizing antibody responses, ADCP also increased in the three-dose (202%), and breakthrough (210%) groups, compared to two-dose vaccination; here as well, the three-dose and breakthrough groups were not significantly different from each other (Fig. 2G).

### Neutralizing antibody responses

Consistent with previous reports, neutralization of live SARS-CoV-2 improved to a greater degree than the observed rise in binding antibody levels (9,17). The geometric mean titers (GMT) showing 50% neutralization of the original SARS-CoV-2 virus (WA1, Wuhan strain) in FRNT assays were 463% and 732% higher for the three-dose and breakthrough groups, respectively, compared to two-dose vaccination, but were not significantly different from each other (Fig. 3A). Among the breakthrough infections, 10 of the 30 participants were infected with the Delta variant. The GMT of the breakthrough group to neutralize the Delta variant increased 948% while the three-dose group increased only 424%, compared to two-dose vaccination. However, the difference between the 3-dose and breakthrough groups did not rise to the level of statistical significance (Fig. 3B). Against Omicron, 25 of 42 (59%) sera in the two-dose group fell below the limit of detection for neutralization, while all three-dose and breakthrough participants showed detectable neutralization, with 1156% and 1810% higher neutralizing GMTs, respectively, which were not significantly different from each other (Fig. 3C).

**Figure 3:**
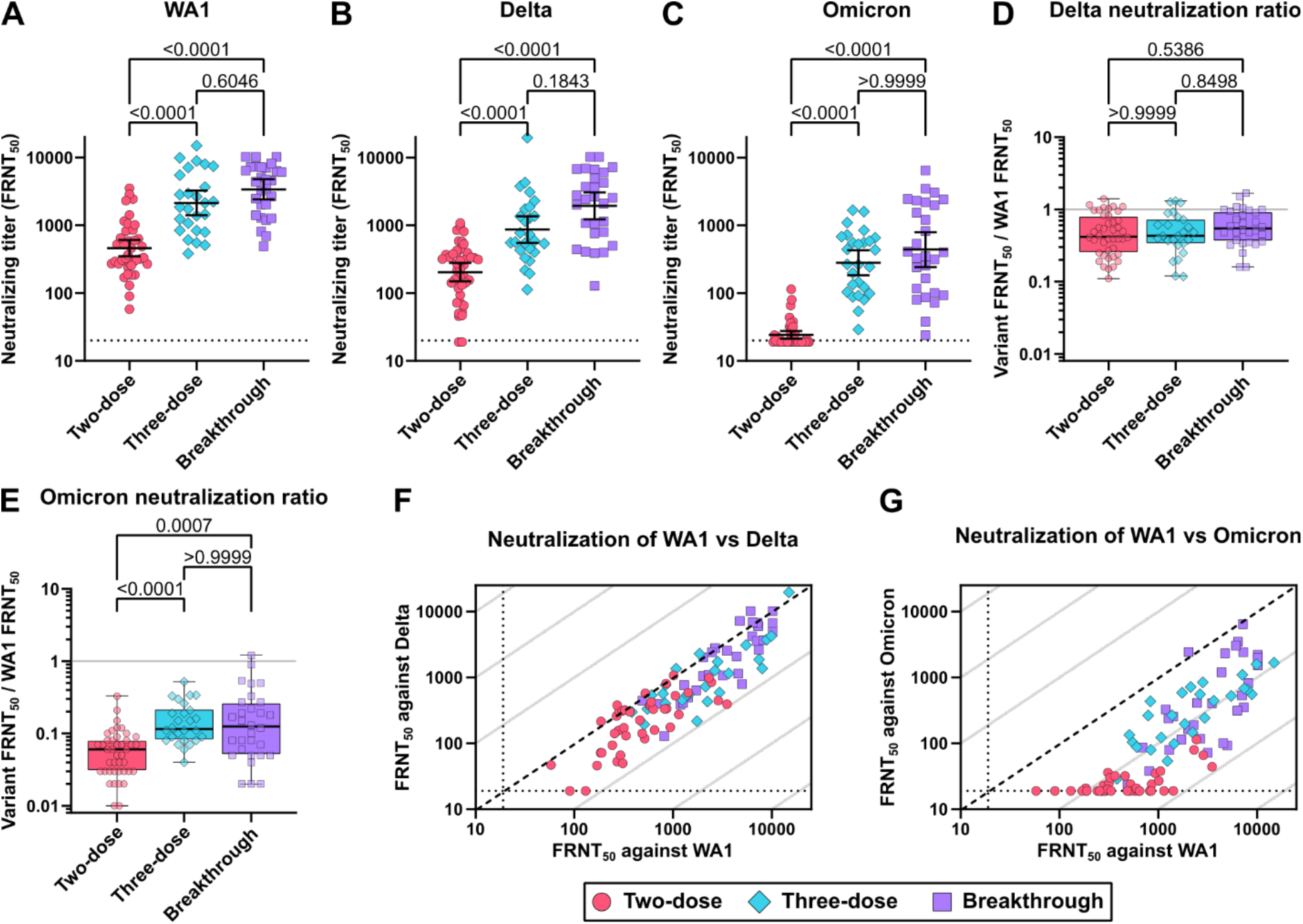
Live SARS-CoV-2 neutralization by two-dose vaccination, three-dose vaccination and breakthrough infection cohorts. (A) Original Wuhan strain WA1, (B) Delta variant, and (C) Omicron variant neutralizing activity determined by 50% focus reduction neutralization test (FRNT_50_). Ratio of (D) Delta and (E) Omicron variant FRNT_50_ over WA1 FRNT_50_. The solid grey lines indicate equal neutralization of variant and WA1. Scatter plots depicting (F) Delta and (G) Omicron variant FRNT_50_ versus WA1 FRNT_50_. The broken lines indicate equal neutralization of variants and WA1, while grey lines signify 10-fold differences. Error bars in A-C indicate the geometric mean and 95% confidence intervals. Box plots in D and E show the median, interquartile range, and full range. P-values in A-E show the results of two-tailed Kruskal-Wallis tests with Dunn’s multiple comparison correction.

### Antibody response quality

The relationship between spike binding antibody level and neutralizing titer gives an indication of the quality of the antibody response by controlling for the total quantity of antibodies present. In all three groups, neutralizing titer correlates strongly with neutralization of WA1 and Delta. However, the correlation is much weaker for the two-dose group for Omicron, largely due to the high proportion of samples below the detection limit (Fig. 4A-4C). We explored this association further by calculating the neutralizing potency index (NPI), as the ratio of live-virus neutralization to spike-specific antibody EC_50_ for WA1, Delta, and Omicron. For WA1, the median NPI was 0.60 for two-dose, 1.10 for three-dose, and 1.91 for breakthrough, showing an increase in the ratio of neutralizing activity to spike binding EC_50_ (Fig. 4D). The median Delta NPI were 0.30, 0.52, and 0.93 and the median Omicron NPI were 0.03, 0.15, and 0.20 for two-dose, three-dose, and breakthrough groups, respectively (Fig. 4E and 4F). For all three viruses, the three-dose and breakthrough groups were found to be significantly increased from the two-dose group, but not significantly different from each other. A similar trend was seen when calculating the NPI using RBD-specific antibody levels instead of those for full-length spike (Fig. 5).

**Figure 4:**
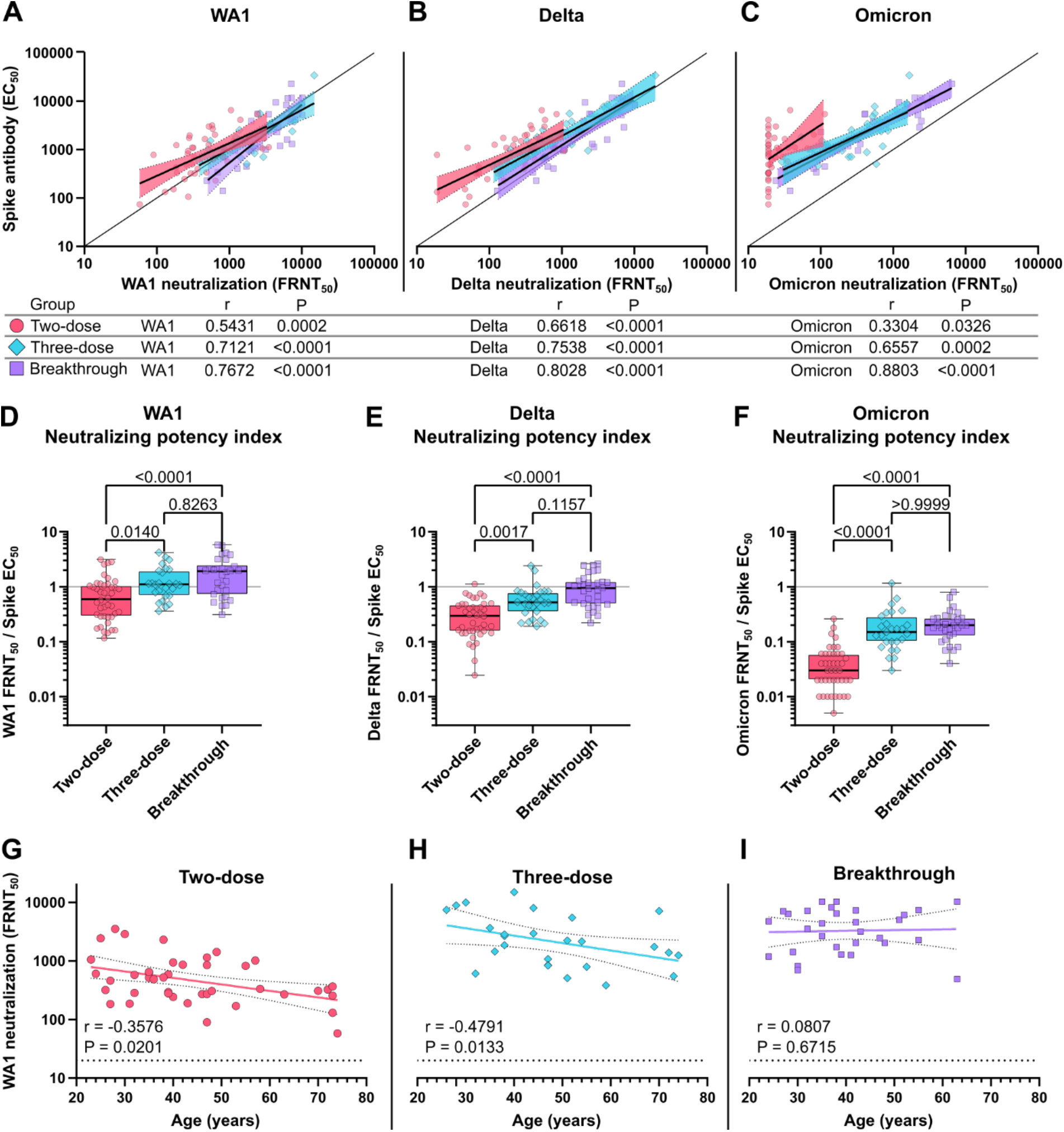
Quality of the neutralizing antibody response to two-dose vaccination, three-dose vaccination and breakthrough infection. Correlation of serum full-length spike-binding antibody EC50 with (A) WA1 FRNT_50_, (B) Delta FRNT_50_, and (C) Omicron FRNT_50_. The solid line indicates equal EC_50_ and FRNT_50_ values. Neutralizing potency indices indicate the ratio of (C) WA1 FRNT_50_, (D) Delta FRNT_50_, and (E) Omicron FRNT_50_ over full-length spike EC_50_. The solid line indicates equal EC_50_ and FRNT_50_ values. Correlation of (G) WA1 FRNT_50_, (H) Delta FRNT_50_, and (D) Omicron FRNT_50_ with age at the time of study enrollment. r in A-C and G-H indicate the Spearman correlation coefficients with corresponding two-tailed P values. Linear best fit-lines with 95% confidence bands were determined by simple linear regression of log transformed EC_50_, FRNT_50_, and non-transformed Age values. Box plots in D-F show the median, interquartile range, and full range. P-values in D-F show the results of two-tailed Kruskal-Wallis tests with Dunn’s multiple comparison correction.

**Figure 5:**
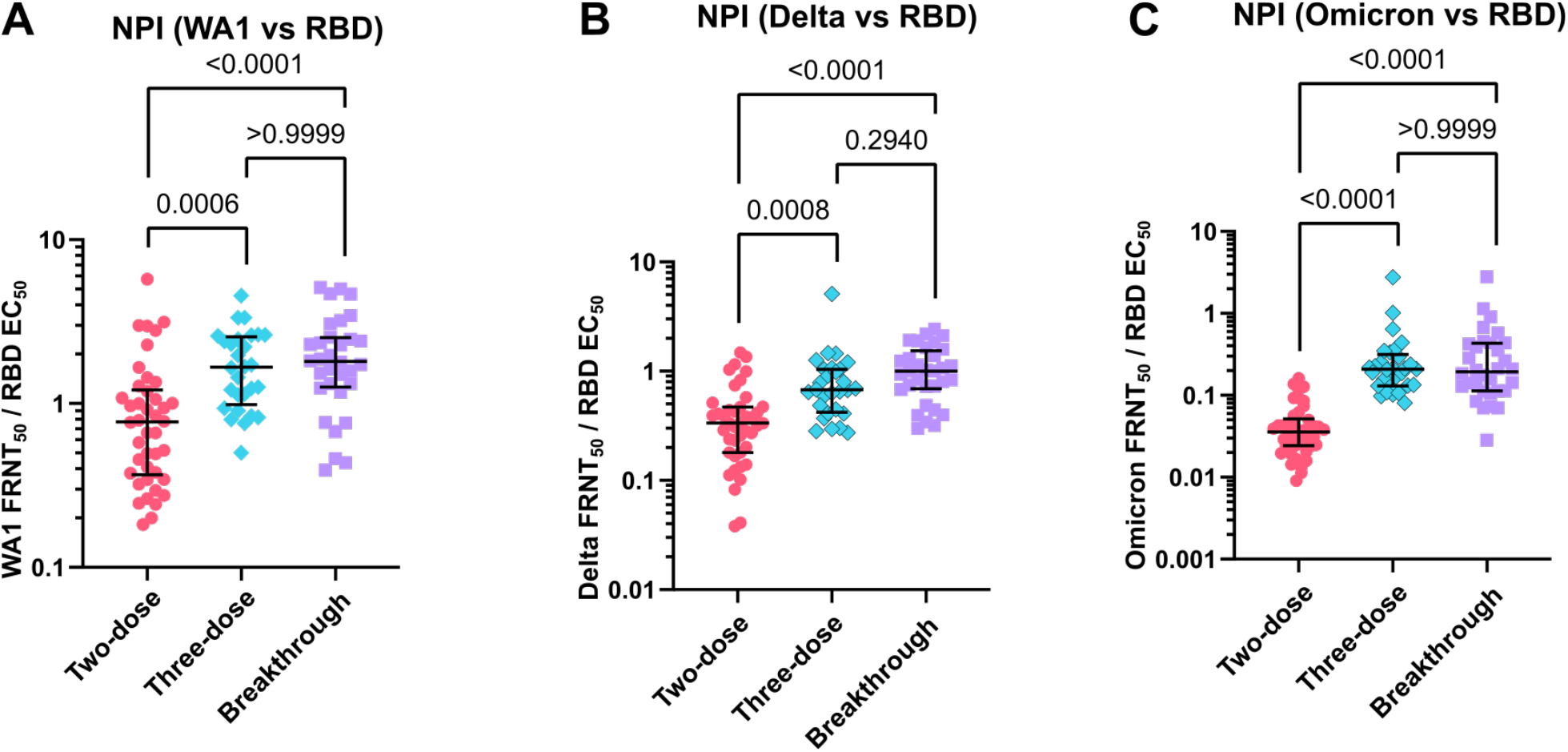
Neutralizing potency indices (NPI) for receptor-binding domain antibodies. NPI indicate the ratio of WA1 50% focus reduction neutralization test (FRNT_50_) (panel A), Delta FRNT_50_, (panel B) and Omicron FRNT_50_ (panel C) to spike receptor binding domain (RBD) EC_50_. Error bars indicate the median and interquartile range. P-values show the results of a two-tailed Kruskal-Wallis test with Dunn’s multiple comparison correction.

### Relative loss of strain-specific neutralizing capacity

To measure the relative loss of neutralizing activity against the Delta and Omicron variants compared to WA1, we calculated the ratio of neutralization for each variant to WA1 neutralization in each participant. For Delta, the median ratio was 0.42 for two-dose, 0.43 for three-dose, and 0.55 for breakthrough, none of which were significantly different (Fig. 3D). Against Omicron, however, the median ratio was 0.06 for two-dose, 0.12 for three-dose, 0.13 for breakthrough, which were significantly higher than two-dose for both the three-dose and breakthrough groups (Fig. 3E). Comparing the neutralization of Delta and Omicron with that of WA1 clearly showed a greater extent of resistance by Omicron, with some individuals displaying nearly a 100-fold reduction in neutralization of Omicron compared to WA1, where grey lines indicate successive 10-fold differences (Fig. 3F and 3G).

### Antibody response vs age and gender

Previous studies have established a negative correlation between antibody response and age among vaccinated individuals (18). Among study participants, we observed a negative correlation between age and WA1 neutralizing titer for the two-dose and also the three-dose groups, but not the breakthrough group (Fig. 4G-3I). We found no difference in neutralizing titer based on gender.

## DISCUSSION

Our data confirm the extent to which Omicron resists neutralization by vaccinated sera, consistent with recent reports (2,3). Importantly, however, we find that both booster vaccination and breakthrough infection enhance neutralizing titers to a similar degree when comparing timepoints approximately 3 months after boosting with 1 month after breakthrough. Despite the reliance of the vaccine on the original SARS-CoV-2 spike protein sequence, we see robust boosting of the Omicron neutralizing response.

While boosting with updated vaccine inserts may ultimately enhance immune responses to new variants such as Omicron, three-dose vaccine regimens and hybrid immune exposures consisting of vaccination and breakthrough infection nevertheless improve the breadth of the humoral response, as seen by improved Delta neutralization and an increased ratio of variant to WA1 neutralizing titers. Further, we see an improvement in the amount of neutralizing activity for a given amount of spike-binding antibody, indicating an improvement in antibody quality. Thus, while two-doses of the currently available mRNA vaccines provide strong protection against symptomatic infection due to the original SARS-CoV-2 and early variants, serological immunity against the Omicron variant is substantially reduced (19), but is restored by booster vaccination or breakthrough infection. A previous study also indicates that hybrid immunity from SARS-CoV-2 infection followed by one or two mRNA vaccine doses provides similar antibody responses to breakthrough infection (9). The similarity seen between immune responses to three-dose regimens and breakthrough infection suggest that the original vaccines continue to provide neutralizing antibody responses that are relevant to the current antigenic landscape, dominated by the Omicron variant. As future variants will emerge from currently circulating strains, the development of vaccines with inserts based on more contemporary variants may further improve the protective efficacy of vaccine-induced immune responses.

Critically, current two-dose vaccine regimens establish a foundation of immunity that is further enhanced through boosting or breakthrough infections, resulting in substantial protection against reinfection, even due to immune-resistant variants such as Omicron. Interestingly, the negative correlation between age and antibody levels seen in vaccines is no longer apparent in hybrid immunity. The enhanced immunity resulting from augmented vaccine regimens and hybrid exposures studied here will apply to an ever-increasing proportion of the world’s population as individuals continue to be vaccinated and exposed to natural infection.

## Data Availability

All data produced in the present study are available upon reasonable request to the authors

## AUTHOR CONTRIBUTIONS

Conceptualization: MEC, FGT, TAB. Recruitment and sample collection: MEC, DS, SDC. Experimental design: MEC, FGT, TAB. Laboratory Analysis: FGT, TAB, GG, SKM. Statistical analysis: TAB. Supervision: MEC, FGT. Manuscript drafting: MEC, TAB. Manuscript review and editing: MEC, TAB, DS, SDC, FGT.

## Contributors

We gratefully recognize the OHSU faculty and staff who participated in this study; the OHSU COVID-19 serology study team and the OHSU occupational health department for recruitment and sample acquisition; Dr. William Messer and research team for sample handling; the OHSU clinical laboratory under the direction of Dr. Donna. Hansel and Dr. Xuan. Qin for SARS-CoV-2 testing and reporting; BEI Resources for providing clinical viral isolates.

